# Healthcare Worker Preparedness for Snakebite Management in Selected Zambian Hospitals: An Exploratory Study

**DOI:** 10.64898/2026.07.17.26358263

**Authors:** Kingford Chimfwembe, Aashna Uppal, Frank Tianyi, Arnold Hamapa, Lydia Hangulu

## Abstract

**Background:** Snakebite envenoming remains a neglected tropical disease and an important public health challenge in Zambia. Effective management of snakebite patients requires healthcare workers who are adequately trained, familiar with treatment protocols, and aware of national management guidelines. However, evidence regarding healthcare worker preparedness for snakebite management in Zambia remains limited. This study explored healthcare worker preparedness for snakebite management in selected hospitals in Zambia.

**Methods:** An exploratory cross-sectional study was conducted in seven hospitals in Zambia between collected between May and July 2025. Twenty-one healthcare workers, comprising senior clinicians, junior clinicians, and nurses, were purposively selected to participate. Data were collected using a structured questionnaire assessing training in snakebite management, clinical exposure to snakebite cases, confidence in management, use of local treatment protocols, and awareness of national snakebite management guidelines. Data were analysed using descriptive statistics and presented as frequencies and percentages.

**Results:** Twenty-one healthcare workers participated in the study. Eight participants (38.1%) reported having received no training in snakebite management, while six (28.6%) reported receiving bedside training. Most participants had recent experience managing snakebite patients, with 66.7% reporting management of at least one snakebite case within the preceding year. Fifteen participants (71.4%) reported being very or exceptionally confident in managing snakebite patients. However, only six participants (28.6%) reported using local snakebite treatment protocols, while eight (38.1%) had seen the latest national snakebite management guidelines. Nearly half of participants reported not using local protocols and had never seen national guidelines.

**Conclusion:** The study identified important gaps in healthcare worker preparedness for snakebite management despite high levels of self-reported confidence. Limited formal training, poor guideline awareness, and low utilization of treatment protocols may affect the quality of snakebite care. Strengthening healthcare worker training and improving dissemination of national management guidelines should be prioritized as part of Zambia’s snakebite control efforts.

## Introduction

Snakebite envenoming remains a major but often overlooked public health challenge, particularly in low- and middle-income countries where access to timely and appropriate treatment is frequently limited [1]. Globally, snakebite envenoming is estimated to affect millions of people annually, resulting in substantial mortality, disability, and socioeconomic consequences [2,3]. In recognition of its significant burden, the World Health Organization [WHO] reinstated snakebite envenoming as a neglected tropical disease and launched a global strategy aimed at reducing snakebite-related deaths and disabilities by 50% by 2030 [1,4].

In Zambia, snakebite envenoming continues to affect rural and peri-urban populations, particularly among farming communities and individuals whose livelihoods place them at increased risk of snake encounters [5,6]. Although the true burden of snakebite remains poorly documented, available evidence suggests that snakebite contributes substantially to morbidity and mortality, particularly in areas where access to healthcare services is limited [7,8]. Delays in seeking healthcare, shortages of antivenom, inadequate surveillance systems, and limited healthcare worker capacity have been identified as important challenges affecting snakebite management in many endemic settings [1,9].

Healthcare workers play a critical role in the diagnosis, treatment, and management of snakebite patients, especially in settings where delayed care can worsen outcomes [10–13]. Appropriate management requires knowledge of snakebite syndromes, recognition of clinical signs of envenoming, use of bedside tests such as the 20-minute whole blood clotting test where appropriate, timely intravenous antivenom administration, and careful management of complications such as bleeding, paralysis, shock, and acute kidney injury [14,15]. Evidence-based protocols also emphasize rapid referral, airway and breathing support, close monitoring, and avoidance of harmful traditional first-aid measure [16]. However, studies conducted in several African and Asian countries have reported deficiencies in healthcare worker training, limited familiarity with treatment guidelines, and inadequate confidence in managing snakebite cases. These deficiencies may contribute to inappropriate clinical management and poor patient outcomes [6,17–20].

Despite growing recognition of snakebite as a public health priority in Zambia, little is known about the preparedness of healthcare workers responsible for managing snakebite patients. Understanding the level of preparedness among healthcare workers is essential for informing capacity-building initiatives, strengthening clinical practice, and improving patient outcomes. This exploratory study therefore assessed healthcare worker preparedness for snakebite management in selected Zambian hospitals, focusing on training, clinical exposure, confidence, protocol utilization, and awareness of national snakebite management guidelines.

## Methods

### Study Design

This exploratory cross-sectional study formed part of a broader assessment of snakebite management and health system preparedness conducted in selected hospitals in Zambia. The study employed a quantitative descriptive approach to explore healthcare worker preparedness for snakebite management.

### Study Setting

The study was conducted in seven hospitals purposively selected from different regions of Zambia: Chipata Central Hospital, Commando Urban Hospital, Gondar Urban Hospital, Kabwe General Hospital, Kitwe Central Hospital, Mpongwe Mission Hospital, and St Dominic’s Mission Hospital. These facilities were selected because they manage snakebite cases and provide varying levels of healthcare services.

### Study Population

The study population comprised healthcare workers providing direct clinical care to patients with snakebite envenoming. This included senior clinicians, junior clinicians, and nurses who were involved in the assessment, diagnosis, treatment, and follow-up management of snakebite cases at the selected health facilities.

### Sampling and Recruitment

A purposive sampling strategy was employed to identify healthcare workers with experience in snakebite management. Three healthcare workers were recruited from each participating hospital, resulting in a total sample of twenty-one participants. Participants were selected based on their involvement in clinical care and their ability to provide information regarding snakebite management practices within their respective facilities.

### Data Collection

Data were collected using a structured questionnaire administered during the study period. Information collected included demographic and professional characteristics, training in snakebite management, clinical experience managing snakebite cases, confidence in snakebite management, use of local treatment protocols, and awareness of national snakebite management guidelines.

### Data Analysis

Data were entered into Microsoft Excel and analysed using descriptive statistics. Frequencies and percentages were calculated to summarise participant characteristics and preparedness indicators. Findings are presented using tables and narrative summaries.

### Ethical Approval

Ethical approval for the study was obtained from the Lusaka Apex Medical University Biomedical Research Ethics Committee (LAMUBREC) (Ref: LAMUBREC No. 0534/13/03/2025; approved on 13 March 2025) and the National Health Research Authority (NHRA), Zambia (Ref: NHRA-2144/17/04/2025; approved on 22 April 2025). Additional administrative permission was obtained from the relevant provincial and district health offices, as well as from the participating health facilities prior to data collection.

## Results

A total of twenty-one healthcare workers participated in the study as shown in Table 1. Participants were drawn equally from seven hospitals, with three participants recruited from each facility. The sample included senior clinicians (33.3%), junior clinicians (33.3%), and nurses (33.3%). Participants possessed varying professional qualifications, including MBChB degrees, diplomas, registered nursing qualifications, and specialist clinical qualifications.

**Table 1.**
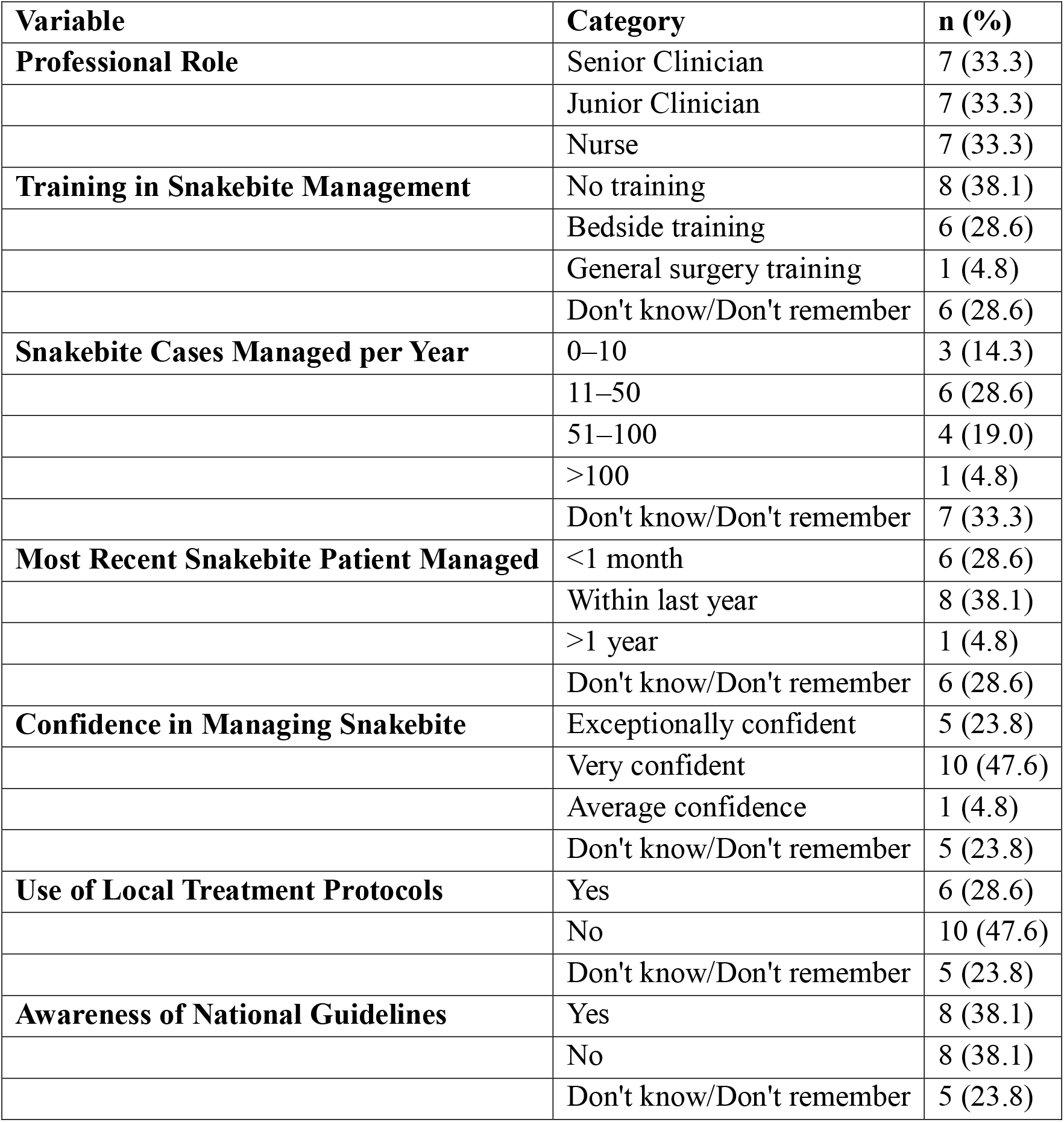
Characteristics and Preparedness of Healthcare Workers for Snakebite Management (N = 21)

Formal training in snakebite management was limited among participants. Eight healthcare workers (38.1%) reported having received no training in snakebite management. Six participants (28.6%) reported learning through bedside training, while only one participant reported receiving training through general surgery. These findings suggest substantial gaps in structured professional development related to snakebite management.

Table 1 demonstrates substantial gaps in formal training, awareness of national guidelines, and utilization of treatment protocols despite relatively high levels of self-reported confidence in snakebite management.

Participants reported varying levels of exposure to snakebite patients. Six participants (28.6%) reported managing between 11 and 50 snakebite cases annually, while four participants (19.0%) reported managing between 51 and 100 cases per year. Most participants had recent experience managing snakebite patients, with 28.6% having managed a case within the previous month and 38.1% having managed a case within the previous year.

Despite limited formal training, participants generally reported high levels of confidence in managing snakebite patients. Ten participants (47.6%) described themselves as very confident, while five (23.8%) reported being exceptionally confident. Only one participant reported average confidence.

The use of local snakebite treatment protocols was inconsistent. Nearly half of participants (47.6%) reported not using local treatment protocols, while only six participants (28.6%) reported using such protocols. Five participants were uncertain whether protocols were available or used within their facilities.

Awareness of national snakebite management guidelines was similarly limited. Eight participants (38.1%) reported having seen the latest national snakebite management guidelines, while an equal proportion reported never having seen them. The remaining participants were uncertain whether they had accessed the guidelines.

## Discussion

The present study explored healthcare worker preparedness for snakebite management in selected hospitals in Zambia and identified important deficiencies in training, protocol utilization, and awareness of national management guidelines. These findings contribute to the growing body of evidence highlighting healthcare system challenges affecting snakebite management in sub-Saharan Africa.

A key finding was the limited access to formal snakebite management training. More than one-third of participants reported never having received any training related to snakebite management [5,9,11,17,21]. Similar findings have been reported in several African countries where healthcare workers frequently encounter snakebite patients but receive little or no formal instruction on diagnosis, antivenom administration, and management of complications [7,22]. Inadequate training has been associated with inappropriate first aid recommendations, delayed administration of antivenom, and inconsistent clinical management practices. These gaps can contribute to incorrect initial treatment, weak adherence to clinical protocols, and poor referral decisions, all of which may worsen patient outcomes. Strengthening pre-service and in-service training for healthcare workers is therefore essential to improve the quality and consistency of snakebite care [13–15,23–25].

Despite these training deficiencies, most participants reported high levels of confidence in managing snakebite patients. This finding suggests a potential mismatch between perceived preparedness and actual preparedness [5,26]. Similar observations have been reported in studies conducted in Nigeria, Kenya, Uganda, and Ghana, where healthcare workers often relied on experiential learning acquired through clinical practice rather than structured professional training. This pattern suggests that snakebite care capacity in these settings may depend heavily on informal workplace learning, which can contribute to variation in knowledge, first aid advice, and treatment decisions [27]. While confidence can enhance decisiveness and clinical responsiveness, excessive confidence in the absence of sufficient evidence-based knowledge may predispose clinicians to inappropriate management decisions and medical error [28–30].

The study also identified poor awareness and utilization of clinical guidance documents. Less than one-third of participants reported using local snakebite treatment protocols, while only a minority had seen the most recent national guidelines [22,31]. Clinical guidelines are essential for standardising patient management, promoting evidence-based practice, and reducing unnecessary variation in care. They are systematically developed to support clinical decisions, translate research into practice, and improve the consistency and quality of care across patients and settings [32–34]. Limited dissemination and implementation of guidelines have been identified as major barriers to effective snakebite management in many low-resource settings [1,5]. The findings suggest that guideline development alone is insufficient unless accompanied by targeted dissemination strategies, training activities, and supportive supervision [7,22].

The relatively high proportion of participants who reported recent exposure to snakebite patients indicates that snakebite remains a clinically relevant condition within participating facilities [3,22]. Consequently, strengthening healthcare worker preparedness has the potential to improve patient outcomes and contribute to broader national efforts aimed at reducing snakebite-related morbidity and mortality. The WHO snakebite roadmap emphasises healthcare worker capacity building as a critical pillar for improving access to safe and effective treatment [1]. The present findings therefore support ongoing efforts to strengthen clinical capacity within Zambia’s health system.

Taken together, the study highlights the need for a comprehensive approach to healthcare worker capacity development. Such an approach should include pre-service training, continuing professional development, routine mentorship, dissemination of national guidelines, and periodic competency assessments [22,31]. These interventions could contribute significantly to improving the quality and consistency of snakebite care in Zambia.

The observed knowledge gaps in this study suggest an urgent need to strengthen dissemination and implementation of national snakebite management guidelines throughout Zambia’s health system. Integrating structured in-service training into continuing professional development programmes and ensuring the routine availability and use of standardised treatment protocols could improve adherence to evidence-based management. Regular mentorship and supportive supervision may further reinforce appropriate clinical practice and enhance the quality of snakebite care.

Future research should employ larger, nationally representative samples and incorporate objective clinical scenarios or direct observation to assess healthcare worker competence beyond self-reported knowledge. Additional implementation research is also needed to identify organisational, contextual and behavioural barriers to guideline uptake and participation in training programmes.

Several limitations should be considered when interpreting the findings. First, the study involved a small purposively selected sample of healthcare workers from seven hospitals and may not be representative of all healthcare workers in Zambia. Second, the study relied on self-reported information, which may be subject to recall bias and social desirability bias. Third, the study assessed perceived preparedness rather than objective clinical competence and therefore could not determine whether reported confidence translated into appropriate clinical practice. Finally, the exploratory design limits the generalisability of findings beyond the participating facilities.

## Conclusion

This exploratory study identified important gaps in healthcare worker preparedness for snakebite management in selected Zambian hospitals. Although participants reported substantial confidence in managing snakebite patients, many lacked formal training and had limited awareness and utilization of national management guidelines and local treatment protocols. These findings suggest opportunities to strengthen healthcare worker capacity and improve the quality of snakebite care through targeted training and enhanced dissemination of evidence-based guidance.

## Data Availability

All data produced in the present study are available upon reasonable request to the authors

## Authors’ contributions

KC: Conceptualization, Methodology, Investigation, Data Curation, Formal Analysis, Project Administration, Writing - Original Draft, Writing - Review & Editing. AU: Methodology, Validation, Writing - Review & Editing. FT: Methodology, Investigation, Validation, Writing - Review & Editing. AH: Writing - Review & Editing. LH: Supervision, Methodology, Writing-Review & Editing. All authors read and approved the final manuscript and agree to be accountable for all aspects of the work.

## Acknowledgements

The authors sincerely acknowledge the Ministry of Health of Zambia at the national, provincial, and district levels for granting permission and facilitating the conduct of this study. We are particularly grateful to the management and staff of the participating health facilities for their cooperation, logistical support, and commitment throughout the study period. We extend our deepest appreciation to all the healthcare workers who generously gave their time to participate in this study. Their willingness to share their knowledge and experiences made this research possible and contributes to ongoing efforts to strengthen the management of snakebite envenoming in Zambia. The authors also thank everyone who provided administrative and technical support during the planning, data collection, and implementation of the study.

